# The impact of medication reconciliation on discrepancies and all-cause readmission among hospitalized patients with chronic kidney disease: A quasi-experimental study

**DOI:** 10.1101/2025.02.10.25321029

**Authors:** Shoroq M. Altawalbeh, Nahlah M. Sallam, Osama Y. Alshogran, Minas Al-Khatib, Mohammad S. Bani Amer, Linda Tahaineh, Abla Albsoul-Younes

## Abstract

**Introduction:** Chronic kidney disease (CKD) with its associated comorbidities and pill burden can expose patients to a heightened risk of drug-related problems, including medication discrepancies.

This study aimed to evaluate the impact of medication reconciliation supplemented with medication review on the number of medication discrepancies at discharge and all-cause readmission among CKD patients.

**Methods:** This was a quasi-experimental trial among adult CKD patients admitted into two major referral hospitals in northern Jordan. Patients in the intervention group received medication reconciliation supplemented with medication review by a clinical pharmacist, while those in the control group received the usual care. The recognized discrepancies were evaluated at admission and at discharge in both groups. Participants were followed for 90-day readmission.

**Results:** Among patients in the intervention group, the average number of discrepancies was 2.5±2.2 per CKD patient. Compared to the control group, the reduction in discrepancy numbers between admission and discharge was higher in the intervention group by 1.66 discrepancies. The likelihood of 90-day readmission was significantly lower in the intervention group (OR=0.41; P=0.002).

**Conclusion:** Supplemented medication reconciliation among CKD patients reveals a favorable impact on medication discrepancies and readmission rates. Optimizing medication management during transitions of care can improve overall health outcomes.

**Impact of findings on practice statements:**

- Activating the role of clinical pharmacists in providing medication reconciliation can decrease medication discrepancies and enhance clinical outcomes particularly in hospitalized CKD patients.
- Designing and implementing an effective interprofessional collaborative approach in the hospital settings might boost the benefits achieved from transition-of-care services.

## Introduction

Medication reconciliation is a healthcare process in which healthcare providers partner with the patient and/or his family/caregiver to ensure that the most accurate and complete medication list is provided (*Canadian Patient Safety Institute (CPSI) and the Institute for Safe Medications Practices Canada (ISMP Canada), Medication Reconciliation in Acute Care Getting Started Kit edition 4, 2017, 69P.*). This service aims to identify and resolve any drug discrepancies throughout the transitions of care in order to improve medication management (*Canadian Patient Safety Institute (CPSI) and the Institute for Safe Medications Practices Canada (ISMP Canada), Medication Reconciliation in Acute Care Getting Started Kit edition 4, 2017, 69P.*). Medication discrepancies are classified as part of drug related problems (DRPs) and are defined as unexplained alterations between medication lists at transitions of care, and may result from poor medication reconciliation (*World Health Organization, Medication safety in transitions of care: technical report, 2019*). Medication discrepancies are considered as one of the leading causes of patient harm and may result in developing adverse drug events (ADEs) and healthcare resource utilization (Abu Farha et al., 2021; Chan et al., 2015; Quelennec et al., 2013; *World Health Organization, Medication safety in transitions of care: technical report, 2019*). Medication reconciliation supplemented with medication review contributed to extra prevention against DRPs (Hammad et al., 2017), and played a significant role in improving clinical and economic outcomes within healthcare institutions (Najafzadeh et al., 2016).

Patients with chronic kidney disease (CKD) are commonly elderly, have multiple comorbidities, require polypharmacy for multi-illness management, receive medications with high-alert risk, have problems with medication adherence, and are prone to higher risk of medication discrepancies (Tesfaye, Peterson, Castelino, McKercher, Jose, Wimmer, et al., 2019). Maintaining an accurate drug list for CKD patients is challenging, as they usually have high comorbidity index which increases the demand for involvement of several prescribers and frequent care transitions (Wilson et al., 2017). Moreover, medication discrepancies have been demonstrated as a modifiable risk for CKD complications and progression (Sakhiya et al., 2020). Proper management of CKD necessitates collaborative efforts of a multidisciplinary healthcare team for optimizing the medication review process (Kelepouris et al., 2023). Previous studies showed the critical role of clinical pharmacist services in achieving positive impact on health outcomes in CKD setting (Al Raiisi et al., 2019). Yet, data about the impact of medication reconciliation on discrepancies and relevant clinical outcomes including re-hospitalization are still insufficient, particularly among inpatients with CKD (Al Raiisi et al., 2019; Whittaker et al., 2018), and warrant further investigations.

### Aim of the study

This study aimed to evaluate the impact of medication reconciliation supplemented with medication review on the number of discrepancies at discharge and the likelihood of all-cause 90-day readmission in CKD admitted patients.

### Ethical approval

This research was approved by the Institutional Review Board (IRB) Committee at KAUH (Ref number: 123/147/2022), and Ministry of Health (Ref number: 13902). This study was performed in line with the principles of the Declaration of Helsinki. An informed consent form was signed by patients upon their approval to participate in the intervention group.

## Methods

### Study Design and Settings

This was a multi-center quasi-experimental clinical study involving patients with stages 2-5 CKD, and was conducted in two major referral hospitals in northern Jordan, namely, KAUH and Princess Basma Hospital (PBH). The study composed of an interventional group, in which patients received a supplemented medication reconciliation service, and a retrospective control group in which patients received the usual care that may or may not include medication reconciliation. Patients in the interventional group were recruited over a 4-month period (February to May 2023) and received medication reconciliation at admission and discharge in addition to continuous medication review during their admissions (supplemented medication reconciliation). In the retrospective control group, patients were identified retrospectively using medical records. Patients who were admitted with CKD diagnosis during the years 2015-2019 were identified as those discharged with ICD-10-CM Diagnosis Code of “N18”. Demographic, clinical, medical and pharmacy data of each patient were retrieved from medical records at admission. Prescriptions 6-months before admission, admission medication lists, medication lists given during hospitalization, prescriptions at discharge and patient medication prescriptions 9-months after discharge were extracted and analyzed to assess discrepancies upon admission and discharge in a methodology adopted from a previous study (Volpi et al., 2021). Figure 1 illustrates the timeframe and outcomes measured in the study groups.

**Figure 1:**
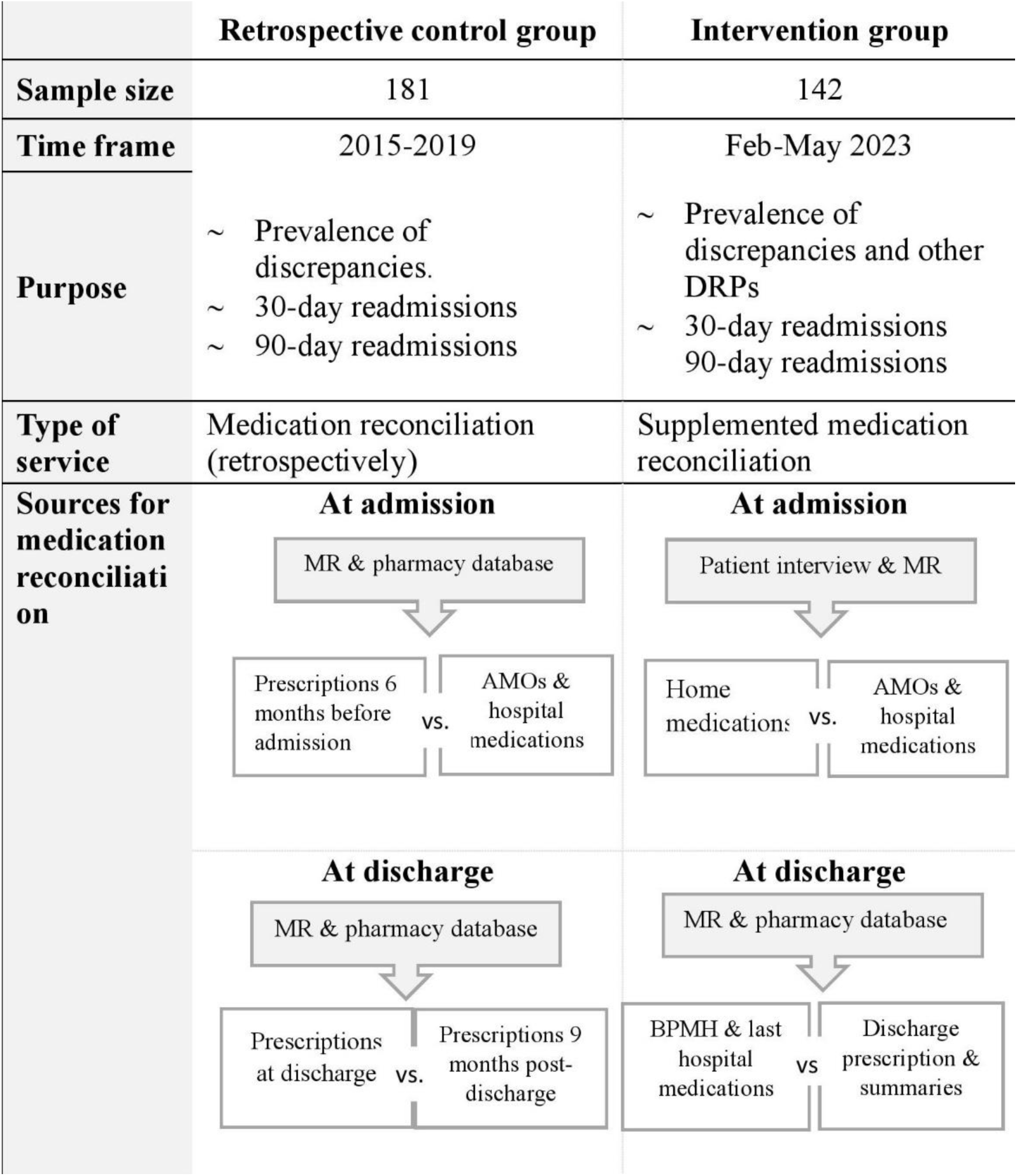
summary of timeline and procedure for each group. AMOs: admission medication orders, MR: medical records, BPMH: best possible medication history, VS: versus.

### Study Outcomes

The primary outcome in this study was the likelihood of all-cause 90-day readmission as extracted from medical records for both study groups. In addition, 30-day readmission was also evaluated. Medication discrepancies were evaluated at admission and at discharge for patients recruited in the intervention group (prospectively) and for patients in the retrospective control group (retrospectively). Classification of discrepancies was made using the Medication Discrepancy Taxonomy (MedTax) system (Almanasreh et al., 2020). Decisions about the response to clinical pharmacist recommendations in the intervention group based on identified discrepancies were categorized into accepted, not accepted, or potentially accepted. Potential accepted status indicated situations in which the clinical pharmacist could not communicate the intervention to the responsible physician/resident on time.

### Inclusion and Exclusion Criteria

Medical records at KAUH and PBH were reviewed to select eligible patients. The study targeted the adult (18 years or older) admitted patients with CKD (stages 2-5 including dialysis), who were using at least three chronic medicines at home, and were hospitalized for at least 48 hours. Patients with kidney transplantation were excluded. Patients who have difficulties in responding to interviews and did not have family member or caregiver on behave, and those who refused to provide a written informed consent were excluded from the intervention group.

### Medication Reconciliation Process Supplemented with Medication Review

Patients in the intervention group received a medication reconciliation model across the transitions of care during the index hospitalization in the internal medicine ward plus medication review for possible DRPs. This service was provided by a trained clinical pharmacist in the internal medicine wards. Eligible subjects received enough information about the purpose and details of the study before signing the informed consent form. At admission, demographic, clinical, and medical information for each enrolled patient were collected from the medical records. Patients or their caregivers were then interviewed to verify demographics, medical history, and pre-admission medications list which was further verified using all other available sources such as bottles, prescriptions, and previous medical records to generate the best possible medication history (BPMH). All comorbidities were rescreened from each patient and Charlson Comorbidity Index (CCI) was estimated (Charlson et al., 1987). At discharge, the best possible discharge medication plan (BPMDP) was created from the BPMH, the last medication list during index hospitalization, and the new medications planned to be started upon discharge. The BPMH, medication reconciliation form, and BPMDP were adapted from the CPSI/ISMP Canada (*Canadian Patient Safety Institute (CPSI) and the Institute for Safe Medications Practices Canada (ISMP Canada), Medication Reconciliation in Acute Care Getting Started Kit edition 4, 2017, 69P.*), with minor modifications. The procedure of the supplemented medication reconciliation for the intervention group was divided into several steps as described in Supplementary Table S1 and depicted in Figure 2.

**Figure 2:**
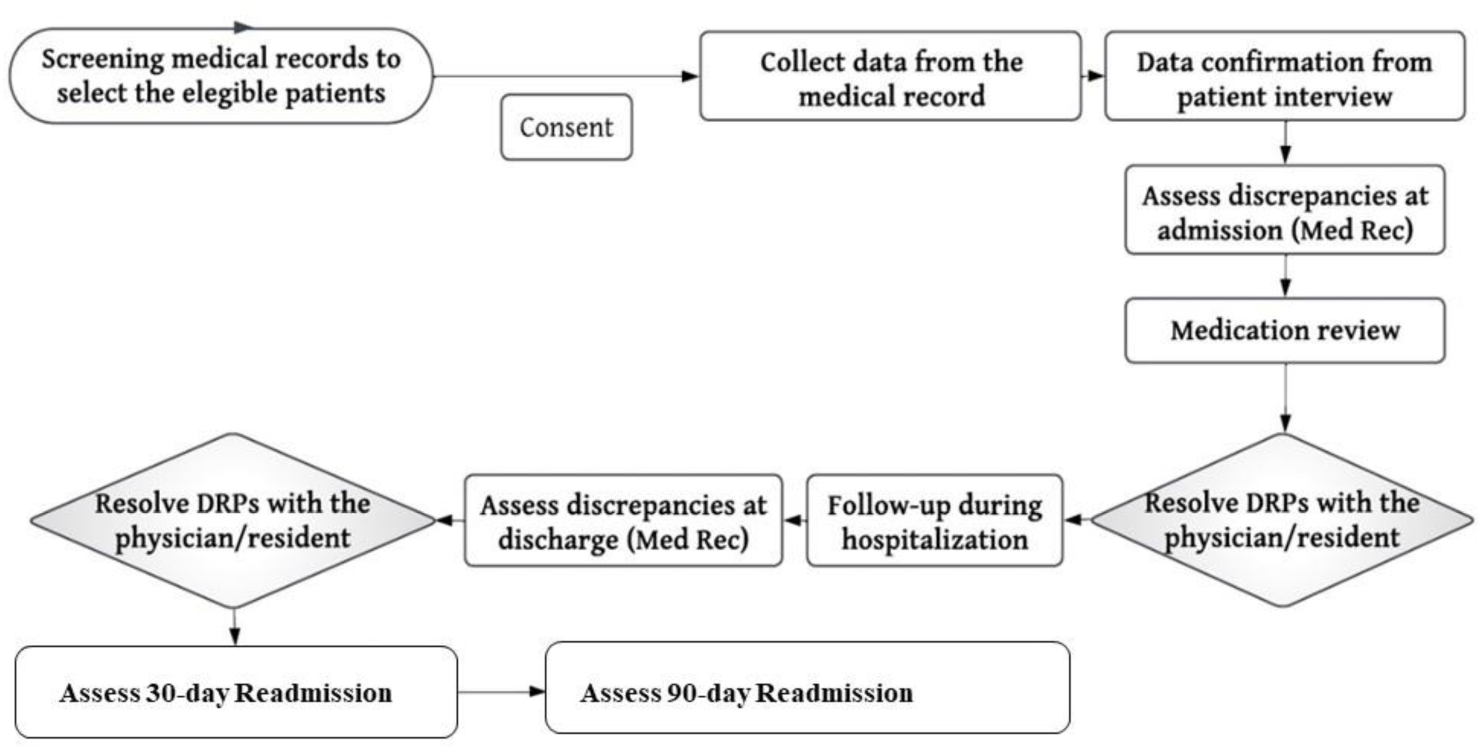
Supplemented medication reconciliation process flow.

### Sample size calculation

Based on a previously reported 90-day readmission proportion of 0.40 among CKD patients (Tesfaye, Peterson, Castelino, McKercher, Jose, Zaidi, et al., 2019), about 264 subjects were required to detect at least 40% decrease in readmission proportion considering a power of 0.8, and a significance level of 0.05.

### Statistical Analysis

Descriptive statistics, such as percentages and arithmetic means with standard deviations were calculated to describe patient characteristics and outcome measures. Unadjusted comparisons between the intervention and control groups were conducted using the t-test, non-parametric (Mann-Whitney) test, or chi-square as appropriate. Compared to the retrospective control group, multi-variable linear regression and logistic regression models were conducted to evaluate the impact of medication reconciliation on reducing the number of discrepancies at discharge and on the likelihood of readmission, respectively. Data were analyzed using Stata version 17 software (StataCorp, 2021, “Stata: Release 17. Statistical Software,” College Station, TX: StataCorp LLC). P-values less than 0.05 were considered statistically significant.

## Results

### Patients’ Socio-demographics and Clinical Characteristics

In total, 323 eligible patients with CKD were included in the study, of whom 142 were recruited prospectively in the intervention period and 181 were included in the retrospective control group. Among eligible patients, 4 patients refused to participate in the intervention group with a response rate of 97.3%. The average patients’ age was 57.16 years old (SD =15.96) in the interventional group, and 62.54 (SD=15.75) in the retrospective cohort group. Patients were distributed across CKD stages 2-5, with a bit more than 50% were on dialysis in both groups. Detailed demographic and clinical characteristics for the interventional and the retrospective control groups are presented in Table 1.

**Table 1:** Demographic characteristics of the recruited sample.

| Variable | Total<br>N = 323 | Intervention<br>N = 142 | Control<br>(Retrospective)<br>N=181 | P<br>value |
| --- | --- | --- | --- | --- |
| Gender n (%) |  |  |  | 0.295 |
| Female | 131 (40.6) | 53 (37.3) | 78(43.09) |  |
| Male | 192 (59.4) | 89 (62.7) | 103(56.91) |  |
| Age, years (M± SD) | 60.2 (16.0) | 57.16<br>(15.96) | 62.54 (15.75) | 0.003 |
| BMI n (%) |  |  |  | 0.504 |
| <18.5 | 8 (2.9) | 6 (4.23) | 2 (1.56) |  |
| 18.5 -24.9 | 85 (31.5) | 41 (28.87) | 44 (43.38) |  |
| 25-29.9 | 78 (28.9) | 42 (29.58) | 36 (28.13) |  |
| > 29.9 | 99 (36.7) | 53 (37.32) | 46 (35.94) |  |
| Marital status |  |  |  | <0.001 |
| Married | 265 (87.2) | 109 (76.76) | 156 (96.3) |  |
| Not married | 39 (12.8) | 33 (23.24) | 6 (3.7) |  |
| Reason for admission |  |  |  | 0.068 |
| AKI on Top of CKD<br>related problems | 127 (39.3) | 42 (29.58) | 85 (46.9) |  |
| CKD/dialysis related<br>problems | 108 (33.4) | 71 (50.0) | 37 (20.4) |  |
| Others | 88 (27.2) | 29 (20.42) | 59 (32.6) |  |
| CKD stage |  |  |  | 0.172 |
| Stage 2 | 3 (0.93) | 2 (1.41) | 1 (0.55) |  |
| Stage 3a | 13 (4.02) | 3 (2.11) | 10 (5.52) |  |
| Stage 3b | 31 (9.6) | 12 (8.45) | 19 (10.5) |  |
| Stage 4 | 85 (26.3) | 32 (22.54) | 53 (29.28) |  |

| Stage 5 | 191 (59.1) | 93 (65.5) | 98 (54.14) |  |
| --- | --- | --- | --- | --- |
| CCI (M ± SD) | 5.84 ± 2.6 | 6.08 ± 2.93 | 5.65± 2.2 | 0.14 |
| LOS (M ± SD) | 9.26 ± 8.8 | 8.9± 8.5 | 9.5± 8.8 | 0.557 |

AKI: acute kidney injury, CKD: chronic kidney disease, CCI: Charlson comorbidity index, LOS: length of stay

### The Prevalence of Discrepancies in CKD patients

In the interventional group, a total of 358 medication discrepancies were identified based on the medication reconciliation at admission and at discharge. The average number of discrepancies was 2.5 ± 2.2 per admission. The most common types of discrepancies were drug omission (70.7%), followed by discrepancies in strength/total daily dose/frequency (14.8%) and unnecessary drug addition (6.4%). Almost half of the interventions were considered as potential to be accepted but were not communicated for physicians (50.5%). Around 38.4% of recommendations were accepted and implemented by physicians/residents.

In the retrospective cohort control group, a total of 471 discrepancies were identified, and the average number of discrepancies was 2.6 ± 2.5 per admission. Drug omission, followed by discrepancies in strength/total daily dose/frequency and therapeutic class substitution were the most common types of discrepancies. Figure S1 (Supplementary Material) depicts discrepancies distribution according to MedTax classification for both the intervention and the retrospective control groups.

### Impact of Medication Reconciliation Intervention on decreasing discrepancy numbers at discharge

The average decrease in discrepancy numbers between admission and discharge in the intervention group was 0.73 compared to -0.91 in the retrospective control group, P<0.001. Adjusting for potential confounders, the decrease in discrepancy numbers between admission and discharge was higher in the intervention group by 1.66 discrepancies compared to the retrospective control group, Table 2.

**Table 2:** Predictors of the decrease in discrepancy numbers between admission and discharge.

| Decrease in discrepancies | Coefficient | 95% CI |  | P value |
| --- | --- | --- | --- | --- |
| Intervention group | 1.66 | 1.07 | 2.26 | <0.001 |
| Age | -0.01 | -0.03 | 0.02 | 0.554 |
| Male gender | 0.02 | -0.56 | 0.61 | 0.936 |
| CCI | 0.07 | -0.09 | 0.22 | 0.411 |
| Dialysis | 0.32 | -0.24 | 0.87 | 0.263 |
| BMI |  |  |  |  |
| Overweight | 0.11 | -0.60 | 0.83 | 0.760 |
| Obese | 0.29 | -0.36 | 0.94 | 0.384 |
| Married | 0.28 | -0.64 | 1.20 | 0.548 |

Analysis was conducted after excluding patients for whom medication reconciliation was not conducted at discharge (49 among the intervention group and 41 among the retrospective cohort group)
Abbreviations: BMI body mass index; CCI: Charlson comorbidity index.

### Impact of Medication Reconciliation Intervention on Hospital Readmission

Compared to the retrospective control group, the interventional group, had significantly lower odds of all-cause 90-day readmissions (OR = 0.41, P =0.002), adjusting for age, gender, CCI, body mass index (BMI), marital status, and receiving dialysis, Table 3. However, no significant impact was seen on 30-day readmission as shown in Table 3.

**Table 3:** Predictors of all-cause readmissions within one month and three months after discharge (adjusted analysis).

| All cause readmission | Odds ratio | 95% CI |  | P value |
| --- | --- | --- | --- | --- |
| 30-day readmission |  |  |  |  |
| Intervention group | 0.62 | 0.32 | 1.19 | 0.150 |
| Age | 1.00 | 0.97 | 1.02 | 0.866 |
| Male gender | 0.74 | 0.39 | 1.39 | 0.352 |
| CCI | 1.14 | 0.97 | 1.34 | 0.104 |
| Dialysis | 1.05 | 0.57 | 1.95 | 0.867 |
| BMI |  |  |  |  |
| Overweight | 1.26 | 0.62 | 2.58 | 0.524 |
| Obese | 0.65 | 0.31 | 1.34 | 0.240 |
| Married | 0.80 | 0.31 | 2.09 | 0.648 |
| 90-day readmission |  |  |  |  |
| Intervention group | 0.41 | 0.23 | 0.72 | 0.002 |
| Age | 1.00 | 0.98 | 1.02 | 0.902 |
| Male gender | 0.69 | 0.40 | 1.19 | 0.180 |
| CCI | 1.03 | 0.89 | 1.19 | 0.699 |
| Dialysis | 1.18 | 0.69 | 2.01 | 0.547 |
| BMI |  |  |  |  |
| Overweight | 1.06 | 0.56 | 2.02 | 0.862 |
| Obese | 0.71 | 0.38 | 1.32 | 0.279 |
| Married | 1.21 | 0.52 | 2.86 | 0.657 |

Patients who died during the index admission were excluded (13 patients; 5 from the intervention group and 8 from the retrospective cohort group)
Abbreviations: BMI body mass index; CCI: Charlson comorbidity index.

## Discussion

The current study showed a positive clinical effect of the medication reconciliation supplemented with medication review on CKD patients’ clinical outcomes. Supplemented medication reconciliation was effective in reducing the number of discrepancies at discharge. The majority of identified discrepancies were due to drug omission at admission and discrepancies in strength, total daily dose, or frequency. Positive impact of medication reconciliation supplemented with medication review was observed on patients’ readmission within 3 months after discharge.

The current study showed that the most frequent type of discrepancies was drug omission followed by discrepancy in strength/total daily dose/frequency. This was in agreement with other studies regarding the drug omission among CKD patients who were not treated with dialysis (Hawley et al., 2019), and in general CKD patients (Song et al., 2021). Chen et al found that the highest percentage of discrepancies linked to incorrect dose/frequency (42%) followed by commissions (38%) and omissions (21%) among hemodialysis patients (Chan et al., 2015). Most of discrepancies occurred at admission which was also reported previously (Tam et al., 2005) as most of discrepancies attributed to uncompleted medication history at admission. In addition, resolving admission discrepancies and DRPs during the admission would definitely have a favorable impact on those at discharge.

The acceptance rate of interventions observed in the current study (38.4%) was relatively comparable or lower against other previous studies conducted in Jordan which reported 38% (Hammour et al., 2022), 54.7% (Salameh et al., 2019), 69.2% (AbuRuz et al., 2013), and 83.9% (Tahaineh & Khasawneh, 2018) in various populations with different outcomes. The relatively low acceptance rate was related in part to the limited timely access to discharge summaries and prescriptions as well as the difficulty in reaching the responsible physician for some cases. Consequently, about half of the DRPs were labeled as potential to be accepted in the current study. Supplemented medication reconciliation was effective in reducing the number of discrepancies at discharge compared to the retrospective CKD admission sample that received the usual care. Resolving discrepancies at admission and the continuous medication review during the admission played a potentially important role in reducing discrepancies from admission to discharge. A previous randomized clinical trial in Jordan evaluated the impact of medication reconciliation on decreasing medication discrepancies after discharge from the surgical ward, and showed a reduction in discrepancy number in both the intervention group and the control group (Hammour et al., 2022). The current study which was conducted particularly in CKD patients showed that decreasing medication discrepancies was significantly higher in the intervention group, which further supports the clinical value of this service in the CKD setting.

Of note, it is well-addressed in previous literature that patients with CKD have markedly higher readmission rate compared to the general populations (Mathew et al., 2015; USRDS, 2021). This study showed a statistically significant difference in all-cause readmissions between the intervention and control groups at 3 months post discharge. This result is highly supported to the literature especially in those with reasonable follow up duration (Chia et al., 2017; Pai et al., 2009). For instance, Chi et al conducted a comprehensive medication review in collaboration approach with nephrologist outpatient clinic for 134 hemodialysis patients who were followed for up to 6 months (Chia et al., 2017), while Pai et al delivered regular medication review to 57 hemodialysis patients up to 2 years (Pai et al., 2009). Additional prospective study involved 50,000 patients undergoing hemodialysis found a significant reduction in rates of readmissions by 0.93 (95% CI, 0.90-0.96) after 30 months of follow-up (Weinhandl et al., 2013). Other published articles on CKD patients found negligible effect on all-cause readmissions (Jasinska-Stroschein, 2022; Song et al., 2021; Tuttle et al., 2018). This may be justified partially by the variability in sample sizes, different interventions provided, and/or variable follow-up periods.

Our study adds to the little existing international literature that evaluated the role of medication reconciliation in CKD patients. Whereas most of these studies focused on end-stage renal disease, the impact of this service is still severely understudied in other CKD stages (Al Raiisi et al., 2019). This study was the first in employing the Quasi experimental design using a retrospective control group in which discrepancies were evaluated and compared with the interventional group without any ethical concerns or contamination. In addition, this study was the first in examining the impact of medication reconciliation on relevant clinical outcomes in admitted patients distributed across various CKD stages. Despite the rigorous efforts, this study has several limitations. The current controlled trial was not randomized which might introduce the risk of selection bias. However, multivariable regression models were applied to control potential confounders. Limited timely access was the main restrictor for the clinical pharmacist. This was related to alternating the days between the two hospitals, and consequently decreasing the availability of the pharmacist at the time of discharge for some patients. In the real-life setting, having a full-time clinical pharmacist is expected to display an effective role and profound effects on patients’ outcomes. In the current study, all-cause readmissions were evaluated without assessing the relation to DRPs. However, we believe that resolving DRPs including discrepancies during hospitalization can have an insightful impact on the general health status, for which all-cause readmission is a valid indicator. Finally, including hospitals in the north of Jordan limits the generalizability of study findings to health practices in the south and middle of Jordan, though not expected to differ widely.

## Conclusions

In conclusion, this study demonstrated that CKD inpatients are very susceptible to DRPs. The current findings suggest that medication reconciliation with medication review service can reduce discrepancy numbers at discharge as well as readmission rates in CKD patients. Multidisciplinary healthcare team that integrates clinical pharmacist is highly recommended for better health outcomes among inpatients with CKD.

## Statements and Declarations

### Data availability statement

The data generated during and/or analyzed during the current study are available from the corresponding author on reasonable request.

### Author contributions

All authors contributed to the study conception and design. Material preparation, data collection and analysis were performed by Shoroq Altawalbeh, Nahlah M. Sallam, Minas Al-Khatib, and Osama Y. Alshogran. The first draft of the manuscript was written by Shoroq Altawalbeh and Nahlah M. Sallam and all authors commented on previous versions of the manuscript. All authors read and approved the final manuscript.

