## Supplementary Table S1, Supplementary Figure S1 for "The impact of medication reconciliation on discrepancies and all-cause readmission among hospitalized patients with chronic kidney disease: A quasi-experimental study"

**Research article**

Supplementary Table S1: Steps of supplemented medication reconciliation process (*Canadian Patient Safety Institute (CPSI) and the Institute for Safe Medications Practices Canada (ISMP Canada), Medication Reconciliation in Acute Care Getting Started Kit edition 4, 2017, 69P.*; *World Health Organization , The High5s Project-Standard Operating Protocol for Medication Reconciliation Standard Operating Protocol Assuring Medication Accuracy at Transitions in Care*, 2014).

| Service | Steps |
| --- | --- |
| Medication reconciliation at admission | 1. BPMH containing all prescribed and nonprescribed medications/supplements was created from patient or family interview and at least one additional reliable source such as medication bottles or boxes, prescriptions, previous medical records. |
|  | 2. The BPMH was compared with the current hospital medication sheet (the admission medication orders) to extract the discrepancies at admission. |
|  | 3. The clinical pharmacist contacted the responsible resident/physician in person or by call to evaluate if the discrepancies were intentional or unintentional and to resolve the unintentional discrepancies. |
| Medication review | 4. The medical information was collected from the medical records (history of present illness, physical assessment data, diagnoses, laboratory information, and vital signs). Some information related to medical history were also gathered during the patient interview. |
|  | 5. A medication review and clinical case analysis were conducted regarding dose adjustments, drug interactions, missing medications, inappropriate medications, unnecessary medications, and monitoring after admission and during hospitalization period to identify any DRPs. |
|  | 6. The clinical pharmacist contacted the responsible resident/physician in person or by call to evaluate and resolve DRPs. |
|  | 7. Daily medication orders review during the index hospital stay was conducted and any new DRPs were communicated to the responsible physician/resident as accessible. |
| Medication reconciliation at discharge | 8. At discharge, the BPMDP was created from the last 24hrs-medication administration list, BPMH, and any new planned discharge medications. |
|  | 9. The BPMDP was compared along with discharge prescription and summaries. Patient education was conducted for willing patients before discharge. |
|  | 10. All identified discrepancies and other DRPs at discharge were discussed with the responsible resident to be resolved, as accessible. |
|  | 11. Decisions about these recommendations and their implementation were categorized into accepted, not accepted, or potential. |

Abbreviations: BPMH: Best Possible Medication History, BPMDP: Best Possible Discharge Medication Plan.

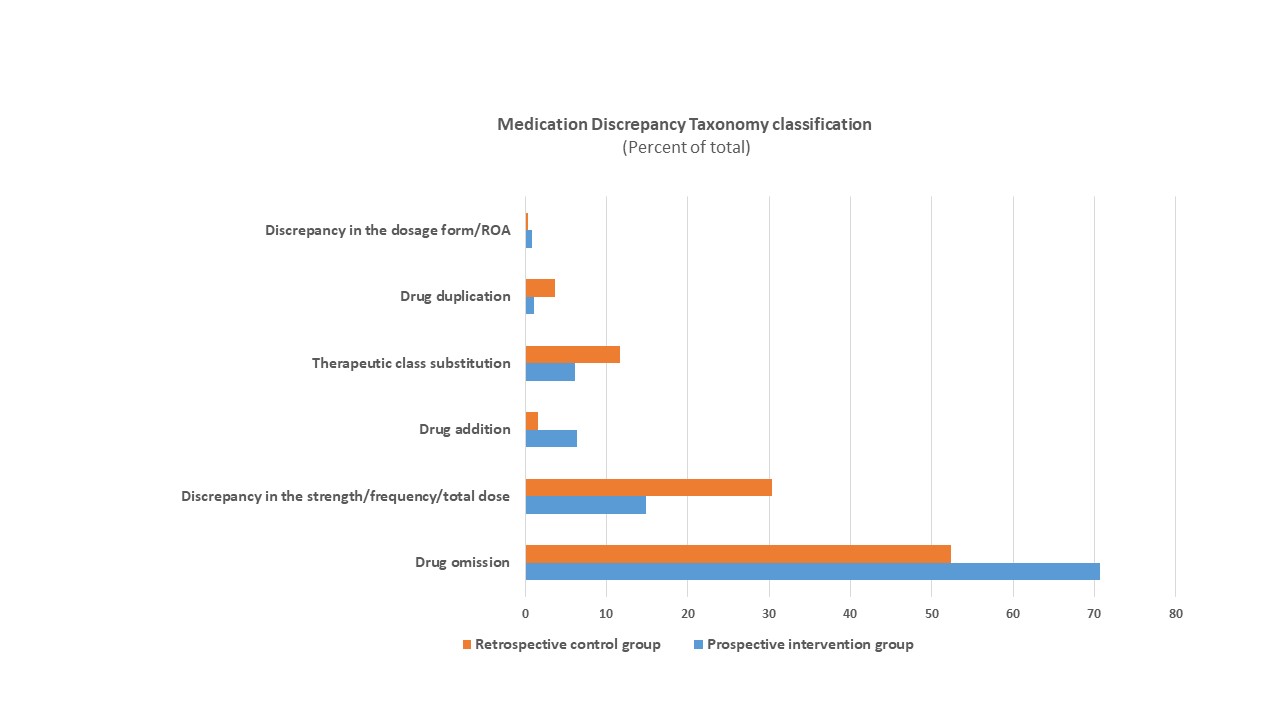

Supplementary Figure S1: Discrepancies distribution among retrospective control and interventional groups according to Medication Discrepancy Taxonomy classification. ROA: route of administration.
